# Effect of high sensitivity C-Reactive Protein on Uric Acid-related Cardiometabolic Risk in Patients with Coronary Artery Disease— A Large Multicenter Prospective Study

**DOI:** 10.1101/2024.06.21.24309325

**Authors:** Ying Song, Weiting Cai, Lin Jiang, Jingjing Xu, Yi Yao, Na Xu, Xiaozeng Wang, Zhenyu Liu, Zheng Zhang, Yongzhen Zhang, Xiaogang Guo, Zhifang Wang, Yingqing Feng, Qingsheng Wang, Jianxin Li, Xueyan Zhao, Jue Chen, Runlin Gao, Lei Song, Yaling Han, Jinqing Yuan

**Affiliations:** Fuwai Hospital, National Center for Cardiovascular Diseases, Chinese Academy of Medical Sciences, Beijing, China; Department of Cardiology, The First Hospital of Lanzhou University, Lanzhou City, China; Department of Cardiology, General Hospital of Northern Theater Command, Shenyang, China; Department of Cardiology, Peking Union Medical College Hospital, Chinese Academy of Medical Sciences and Peking Union Medical College, Beijing, China; Department of Cardiology, Peking University Third Hospital, Beijing, China; Department of Cardiology, The First Affiliated Hospital, Zhejiang University School of Medicine, Hangzhou, China; Department of Cardiology, Xinxiang central Hospital, Xinxiang, China; Department of Cardiology, Guangdong Provincial People’s Hospital, Guangzhou, China; Department of Cardiology, The First Hospital of QinHuangDao, Qinhuangdao, China

**Keywords:** Coronary artery disease, systemic inflammation, high-sensitivity C-Reactive Protein, Uric Acid

## Abstract

**Aims:** Although serum uric acid (SUA) is a risk factor for cardiometabolic outcome, but it remains unclear which patients with coronary artery disease (CAD) benefit the most from SUA lowering therapy (ULT). The association of SUA level, systemic inflammation and cardiometabolic risk is still unclear. The current study is aimed to examine whether SUA-associated cardiometabolic risk is modulated by systemic inflammation in CAD patients.

**Methods and Results:** A total of 16,598 CAD patients with baseline high-sensitivity C-Reactive Protein (hsCRP) and SUA available were included. Baseline and follow-up data were collected. The primary endpoint was major adverse cardiovascular and cerebrovascular events (MACCE), including death, myocardial infarction and stroke. In patients with hsCRP ≥2 mg/L, increasing quintiles of SUA were significantly associated with increased rates of 2-year MACCE (adjusted p < 0.001 for trend, p = 0.037 for interaction). Each unit increase in SUA levels was associated with a 11.3% increased risk of MACCE (adjusted p < 0.001, p = 0.002 for interaction). However, in patients with hsCRP < 2mg/L, increasing quintiles of SUA were not associated with increased MACCE (adjusted p = 0.120).

**Conclusion:** Elevated SUA levels are related to MACCE when hsCRP levels are 2 mg/L or more but not less than 2mg/L. This finding suggests a potential benefit of combined ULT and anti-inflammation therapy in patients with hyperuricemia and greater systemic inflammation.

## Introduction

Coronary artery disease (CAD) still expands threats to global health and remains one of the leading cause of morbidity and mortality worldwide [1]. Despite standard secondary prevention and guideline recommended therapy, CAD patients are still confronted with risk of recurrent adverse cardiovascular and cerebrovascular events. Although intensive management of traditional risk factors such as LDL-C, hypertension, diabetes mellitus and smoking have made progress on reducing ischemic events, novel risk factors remains to be recognized to further improve clinical prognosis.

As the prevalence of hyperuricemia has been increasing worldwide [2, 3], several studies have reported that hyperuricemia was associated with increased CAD risk and adverse cardiometabolic outcomes in CAD patients [4–8]. Serum uric acid (SUA) level is used to diagnose hyperuricemia and to monitor and guide urate-lowering therapy (ULT). The pathogenesis of increased SUA level and atherosclerosis was not fully interpretated. High-sensitivity C-reactive protein (hsCRP) is a widely used biomarker for systematic inflammation and a novel anti-inflammation therapy target for atherosclerotic disease [10, 11]. Interestingly, previous study reported that inflammation played important roles in various signal pathways for SUA activated atherosclerosis [9] and uric acid induced hs-CRP express in vascular cell proliferation[12]. However, whether hs-CRP could modulate SUA-associated cardiometabolic risk was still unclear. Accordingly, we tested the hypothesis that SUA-associated adverse cardiovascular and cerebrovascular outcome would be significantly modulated by systemic inflammation in a large, multicenter, contemporary cohort of CAD patients.

## Method

### Study population

We conducted a post-hoc analysis of data from the **PR**ospective **O**bservational **M**ulticenter cohort for **I**schemic and h**E**morrhage risk in coronary artery disease patients (the PROMISE cohort). Briefly, the PROMISE cohort was a multicenter prospective cohort, enrolled 18,701 patients with coronary artery disease at 9 centers in China from January, 2015 to March, 2019. Inclusion criteria included patients of at least 18 years old, diagnosis of CAD, indication for at least one antiplatelet drug. Exclusion criteria were a life expectancy of fewer than 6 months and current participation in another interventional clinical trial. The selection of a treatment strategy was dependent on clinician and patient preference in accordance with contemporaneous guidelines [13]. Coronary angiography and intervention were performed by experienced coronary interventional cardiologists. The decision for percutaneous coronary intervention (PCI) or coronary artery bypass grafting (CABG) was made by a heart team composed of experienced cardiologists and surgeons.

### Biomarker Assessment

Blood samples were taken by direct vein puncture with 24 hours after admission. The samples were collected into general vacuum tube without anticoagulation. SUA was measured by Uricase Colorimetric Assay and the upper reference limit (URL) was 1.33mg/dl (to convert millimoles per liter to milligram per deciliter, divided by 59.5). And hs-CPR was measured by immunoturbidimetry and the URL was 3mg/L.

### Clinical End Points

All patients were evaluated by clinic visit or by phone at 1, 6, 12 and 24 months. The ischemic and bleeding endpoints were recorded at follow up. The primary endpoint was major adverse cardiovascular and cerebrovascular events (MACCE), a composite of death, myocardial infarction (MI) and stroke. The secondary endpoint were death, MI and stroke. MI was diagnosed in accordance with the contemporaneous Universal Definition of Myocardial Infarction [14, 15]. Strokes included ischemic and hemorrhagic strokes according to the World Health Organization classification of diseases. [16] Investigator training, blinded questionnaire filling, and telephone recording were performed to obtain high-quality data. All endpoints were adjudicated centrally by 2 independent cardiologists, and disagreement was resolved by consensus.

### Statistical Analysis

Continuous variables are reported as mean and SD values for normally distributed variables or median and interquartile range values for non-normally distributed variables. Categorical variables are reported as frequency and percentage. Multivariable Cox stepwise hazards regression modeling was used to assess the relationship between MACCE and SUA, including interactions, while adjusting for important covariates. Hazard ratios and 95% CIs are reported. Tests of trend were performed across the SUA quintiles. Kaplan-Meier curves illustrate the incidence of MACCE during follow up. The number at risk at each 200 days increment is given under each plot. Missing values were imputed using the median for continuous variables or the mode for categorical variables. All P values were from 2-tailed tests and results were deemed statistically significant at P < .05. Analysis was performed using SPSS, version 24.0 (IBM Corp., Armonk, NY, USA). Figures were created using GraphPad Prism, version9.0.0(86) (GraphPad Software, LCC).

## Results

### Patients and baseline characteristics

The present study included patients with both SUA and hsCRP available within 24 hours after admission. As such, 16,598 of the enrolled 18,701 patients in PROMISED cohort met the criteria and were included in the present analysis.

Table 1 describes the baseline clinical and biomarker characteristics of the current population stratified according to achieved hsCRP levels (<2mg/L vs. ≥2mg/L). The average age of the present population was 60±10 years old and most patients were male (12410[67.5%]), 5389(32.5%) patients had diabetes mellitus, 10857(65.4%) and 12675(76.4%) had hypertension and hyperlipidemia respectively. 15903(95.8%) patients received aspirin, 15203(91.6%) received P2Y12 receptor antagonist, 15763(95.0%) received statins. The average levels of SUA was 5.79±1.65mg/dl and hsCRP was 3.87±5.61 mg/L. When baseline characteristics were stratified by hsCRP levels, patients with hsCRP ≥ 2mg/L had higher body mass index, low-density lipoprotein cholesterol, Lipoprotein(a) and baseline Synergy Between Percutaneous Coronary Intervention with Taxus and Cardiac Surgery (SYNTAX) score. Patients with hsCRP ≥ 2mg/L had more comorbidity of hypertension, chronic kidney disease, prior stroke, smoking history, heart failure of reduced ejection fraction (HFrEF) and acute myocardial infarction.

**Table 1.** Baseline patient characteristics according to achieved hsCRP levels.

| **Variable** | **hsCRP < 2 mg/L**<br>**(n=8533)** | **hsCRP ≥ 2 mg/L**<br>**(n=8065)** | **P value** |
| --- | --- | --- | --- |
| **Age, mean (SD), y** | 61±10 | 61±11 | <0.001 |
| **Male sex** | 6423(75.3) | 5987(74.2) | 0.124 |
| **BMI mean (SD) (kg/m2)** | 25.6±3.2 | 26.2±3.5 | 0.001 |
| **AMI** | 1347(15.8) | 3976(49.3) | <0.001 |
| **Hypertension** | 5478(64.2) | 5379(66.7) | 0.001 |
| **Diabetes Mellitus** | 2765(32.4) | 2624(32.5) | 0.868 |
| **Hyperlipidemia** | 6595(77.3) | 6080(75.4) | 0.004 |
| **COPD** | 121(1.4) | 145(1.8) | 0.055 |
| **Chronic Kidney disease** | 115(1.3) | 186(2.3 ) | <0.001 |
| **PVD** | 508(6.0) | 467(5.8) | 0.668 |
| **Prior stroke** | 1264(14.8) | 1323(16.4) | 0.005 |
| **Prior myocardial infarction** | 1651(19.3) | 1359(16.9) | <0.001 |
| **Prior PCI** | 2392(28.0) | 1818(22.5) | <0.001 |
| **Prior CABG** | 226(2.6) | 170(2.1) | 0.025 |
| **Smoking history** | 4748(55.6) | 4912(60.9) | <0.001 |
| **HFrEF** | 210(2.5) | 411(5.1) | <0.001 |
| **Revascularization** | 6086(71.3) | 6564(81.4) | <0.001 |
| **PCI** | 5750(67.4) | 6241(77.4) | <0.001 |
| **CABG** | 395(4.6) | 384(4.8) | 0.714 |
| **Baseline SYNTAX score** | 12.4±8.8 | 14.5±9.3 | <0.001 |
| **Residual SYNTAX score** | 7.0±8.1 | 7.6±8.8 | <0.001 |
| **hsCRP, mg/L** | 0.9±0.6 | 7.0±6.8 | <0.001 |
| **SUA, mg/dl** | 5.7±1.4 | 5.9±1.7 | <0.001 |
| **LDL-C, mmol/L** | 2.3±0.8 | 2.6±0.9 | <0.001 |
| **Lpa, mg/L** | 295±309 | 317±308 | <0.001 |
| **Medications** |  |  |  |
| **Aspirin** | 8176(96.3) | 7727(96.2) | 0.683 |
| **Clopidogrel** | 5929(69.8) | 5638(70.2) | 0.634 |
| **Ticagrelor** | 1634(19.2) | 2002(24.9) | <0.001 |
| **CCB** | 3914(46.1) | 3039(37.8) | <0.001 |
| **ACEI/ARB** | 4219(49.7) | 4577(57.0) | <0.001 |
| **Statins** | 8123(95.7) | 7640(95.1) | 0.075 |

hsCRP: high-sensitivity C-Reactive protein; BMI: body mass index; AMI: acute myocardial infarction; COPD: Chronic obstructive pulmonary disease; PVD: peripheral vascular diseases; PCI: percutaneous coronary intervention; CABG: coronary artery bypass graft; HFrEF: heart failure with reduced ejection fraction; SYNTAX score: the Synergy Between Percutaneous Coronary Intervention with Taxus and Cardiac Surgery score; SUA: serum uric acid; LDL-c: low density lipoprotein C; Lp(a): lipoprotein a; CCB: calcium channel blocker; ACEI/ARB: angiotensin-converting enzyme inhibitor/ angiotensin receptor blocker.

### Clinical outcomes

A total of 16,164(97.4%) patients completed 2-year follow up. During 2-year follow up, 459(2.8%) patients had death, 325(2.0%) had cardiac death, 235(1.4%) had myocardial infarction, 382(2.3%) had stroke and 1795(10.8%) had MACCE. Table 2 shows the 2-year cardiovascular and cerebrovascular events according to sCRP levels. Patients with hsCRP levels ≥ 2 mg/L experienced higher risk of 2-year death (4.3% vs. 1.4%, p < 0.001), cardiac death (3.1% vs. 0.9%, p < 0.001), myocardial infarction (1.9% vs. 1.0%, p < 0.001), stroke (2.7% vs. 2.0%, p = 0.004), and MACCE (7.5% vs. 3.9%, p < 0.001) than patients with hsCRP <2 mg/L.

**Table 2.** The MACCE events rates in the overall hsCRP and achieved hsCRP levels population.

|  | **Overall hsCRP population**<br>**(n=16598)** | **hsCRP < 2 mg/L**<br>**(n=8533)** | **hsCRP ≥ 2 mg/L**<br>**(n=8065)** | **P** |
| --- | --- | --- | --- | --- |
| **Death** | 459(2.8) | 115(1.4) | 344(4.3) | <0.001 |
| **Cardiac Death** | 325(2.0) | 78(0.9) | 247(3.1) | <0.001 |
| **Myocardial Infarction** | 235(1.4) | 85(1.0) | 150(1.9) | <0.001 |
| **Stroke** | 382(2.3) | 168(2.0) | 214(2.7) | 0.004 |
| **MACCE** | 935(5.8) | 329(3.9) | 606(7.5) | <0.001 |

Figure 1 shows the incidence of first MACCE stratified according to SUA quintiles in the setting of hsCRP <2 mg/L (Figure 1a) or ≥2 mg/L (Figure 1b) over 2-year follow up. Only in those with hsCRP ≥2 mg/L, increasing quintiles of SUA levels were associated with a higher incidence of 2 years MACCE (P < .001 for trend). However, in those with hsCRP levels < 2 mg/L, increasing quintiles of SUA levels were not associated with a higher incidence of MACCE (P = 0.403 for trend).

**Figure 1.**
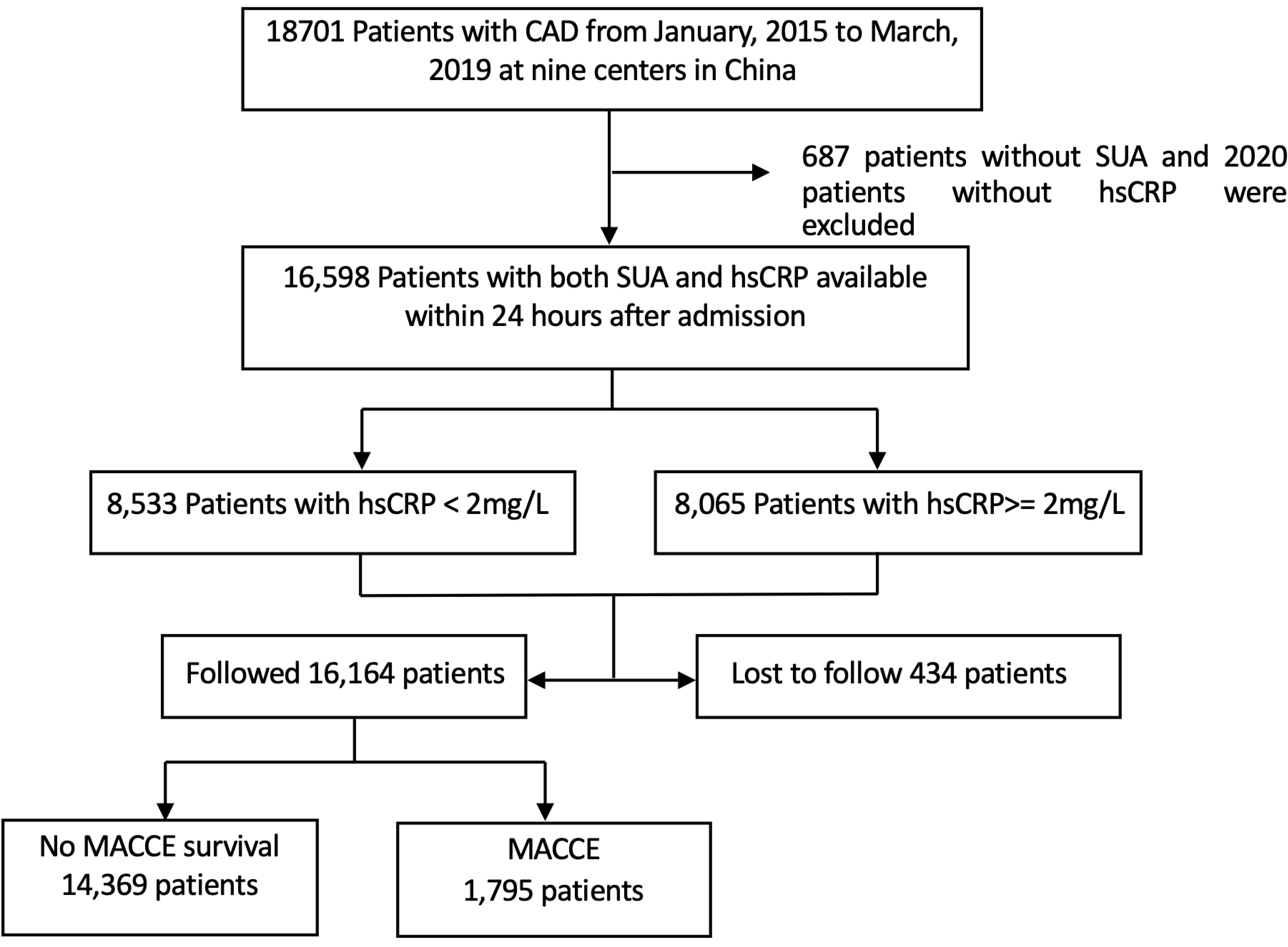
A flow chart for subject selection. CAD, coronary artery disease; SUA, serum uric acid; hsCRP, high-sensitivity C-reactive protein; MACCE, major adverse cardiovascular and cerebrovascular disease.

**Figure 2.**
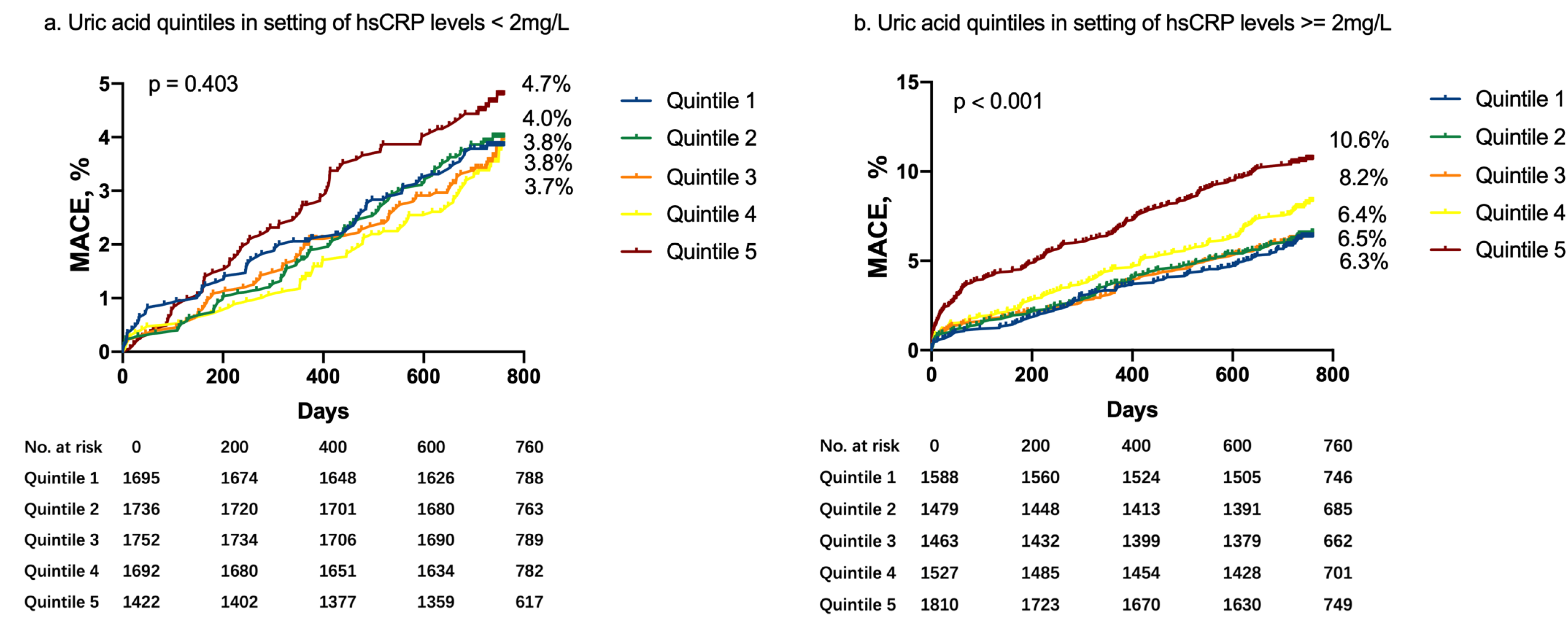
Kaplan-Meier curves of MACCE according to SUA quintiles in the setting of achieved hsCRP levels of less than 2 vs. 2mg/dl or more. SUA, serum uric acid; hsCRP, high-sensitivity C-reactive protein; MACCE, major adverse cardiovascular and cerebrovascular disease.

Table 3 shows univariable and multivariable adjusted relationships between MACCE stratified according to hsCRP and SUA levels. In the overall population, compared to hsCRP level < 2 mg/L, higher hsCRP level (≥ 2 mg/L) was significantly associated with higher risk of MACCE (HR = 1.45, 95%CI: 1.25-1.68, p < 0.001). And the continuous hsCRP levels were also associated with MACCE (HR = 1.02, 95%CI: 1.01-1.02, p < 0.001). In the overall population, SUA levels were associated with MACCE (HR = 1.06, 95%CI: 1.04-1.06 p < 0.001). However, after being stratified by hsCRP of 2mg/L , SUA levels were significantly associated with MACCE only when hsCRP level ≥ 2 mg/L and each unit increase of SUA levels was associated with 11.3% increased risk of MACCE (HR = 1.13, 95%CI: 1.08-1.18, p <0.001). In patient with hsCRP level < 2 mg/L, SUA levels were not significantly associated with higher MACCE (HR = 1.04, 95%CI: 0.99-1.08, p = 0.120). There was a significant interaction for MACCE between hsCRP dichotomy and SUA levels (P = 0.002 for interaction).

**Table 3.** Univariable and multivariable analysis of MACCE according to achieved hsCRP Levels and Continuous SUA.

|  | Crude HR (95%CI) | p | Adjusted<sup>a,b</sup> HR (95%CI) | p |
| --- | --- | --- | --- | --- |
| **Overall hsCRP population** |  |  |  |  |
| < 2 mg/L | reference | — | reference | — |
| ≥ 2mg/L | 1.99(1.74-2.27) | <0.001 | 1.45(1.25-1.68) | <0.001 |
| hsCRP, mg/L | 1.02(1.02-1.03) | <0.001 | 1.02(1.01-1.02) | <0.001 |
| **Overall SUA population** |  |  |  |  |
| SUA, mg/dl | 1.08(1.06-1.09) | <0.001 | 1.06(1.04-1.06) | <0.001 |
| **hsCRP ≥ 2 mg/L** |  |  |  |  |
| SUA, mg/dl | 1.16(1.11-1.21) | <0.001 | 1.13(1.08-1.18) | <0.001 |
| **hsCRP < 2 mg/L** |  |  |  |  |
| SUA, mg/dl | 1.04(0.99-1.09) | 0.114 | 1.04(0.99-1.08) | 0.120 |
| a. multivariable factors: sex, age, acute myocardial infarction, previous myocardial infarction, hypertension, heart failure with reduced ejection fraction, smoking, chronic kidney disease, percutaneous coronary intervention |  |  |  |  |
| b. P = 0.002 for interaction between continuous SUA and hsCRP 2 mg/L or more vs less than 2 mg/L for MACCE |  |  |  |  |
| hsCRP: high-sensitivity C-reactive protein; SUA: serum uric acid |  |  |  |  |

Figure 3 shows the restricted cubic splines to flexibly model and visualize the adjusted relation of predicted SUA level with MACCE. In the whole population, the risk of MACCE was linearly increased with SUA levels and after reaching SUA level of 5.63mg/dl, MACCE risk was significantly higher with increased SUA levels (p < 0.001) (Figure 3A). In patients with hsCRP level < 2 mg/L, the curve was relatively flat when SUA levels were below 5.58mg/dl and there was no significant association between MACCE risk and SUA levels (p = 0.12) (Figure 3B). However, in population with hsCRP level ≥ 2 mg/L, the plot showed a sharp increase in the risk of MACCE, especially within the higher range of predicted SUA level (above 5.68mg/dl). The increased MACCE risk became significantly related to predicted SUA levels. (p < 0.001) (Figure 3C).

**Figure 3.**
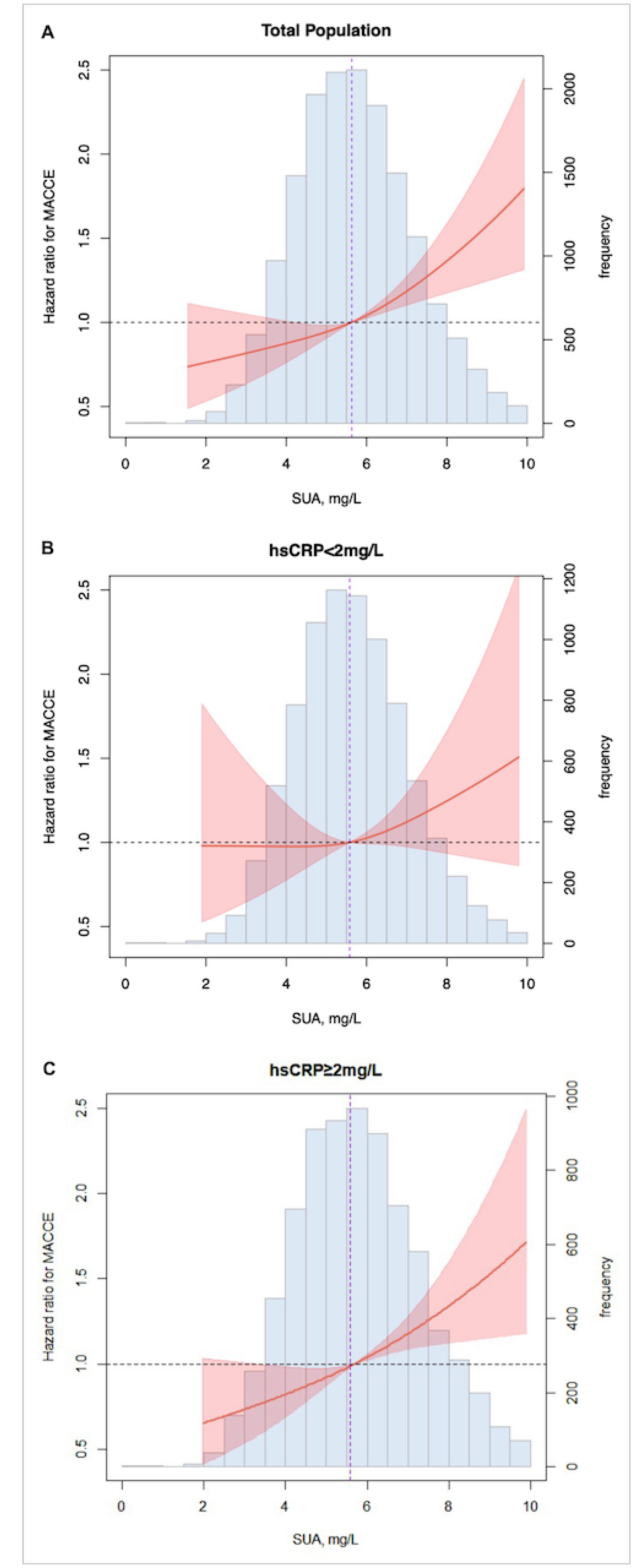
Association of adjusted SUA levels with MACCE in the whole population(3A), hsCRP<2mg/dl(3B) and hsCRP≥2mg/dl(3C) We used continuous SUA levels to analysis the association with MACCE risk in adjusted models. The multivariable model adjusted for sex, age, acute myocardial infarction, previous myocardial infarction, hypertension, heart failure with reduced ejection fraction, smoking, chronic kidney disease, percutaneous coronary intervention. The lines represent adjusted hazard ratios (solid lines) and 95% confidence intervals (shaded areas). The dashed vertical lines in 3A, 3B and 3C were separately stood for SUA levels of 5.63mg/dl, 5.58md/dl and 5.68mg/dl. SUA, serum uric acid; hsCRP, high-sensitivity C-reactive protein; MACCE, major adverse cardiovascular and cerebrovascular disease.

Table 4 describes the associations between hsCRP and SUA quintiles. After fully adjusted multivariable analysis, hsCRP≥2mg/L subgroup, SUA quintiles were associated with higher MACCE(p for trend < 0.001), and high SUA quintiles(Q3 [6.05-6.98mg/dl]and Q4[6.99-61.65mg/dl]) were significantly associated with a greater risk of MACCE. However, when hsCRP < 2 mg/L, high SUA quintiles were not associated with higher risk of MACCE (p for trend = 0.278). There was a significant interaction for MACCE between SUA quintiles and hsCRP dichotomy (P = 0.037 for interaction).

**Table 4.**
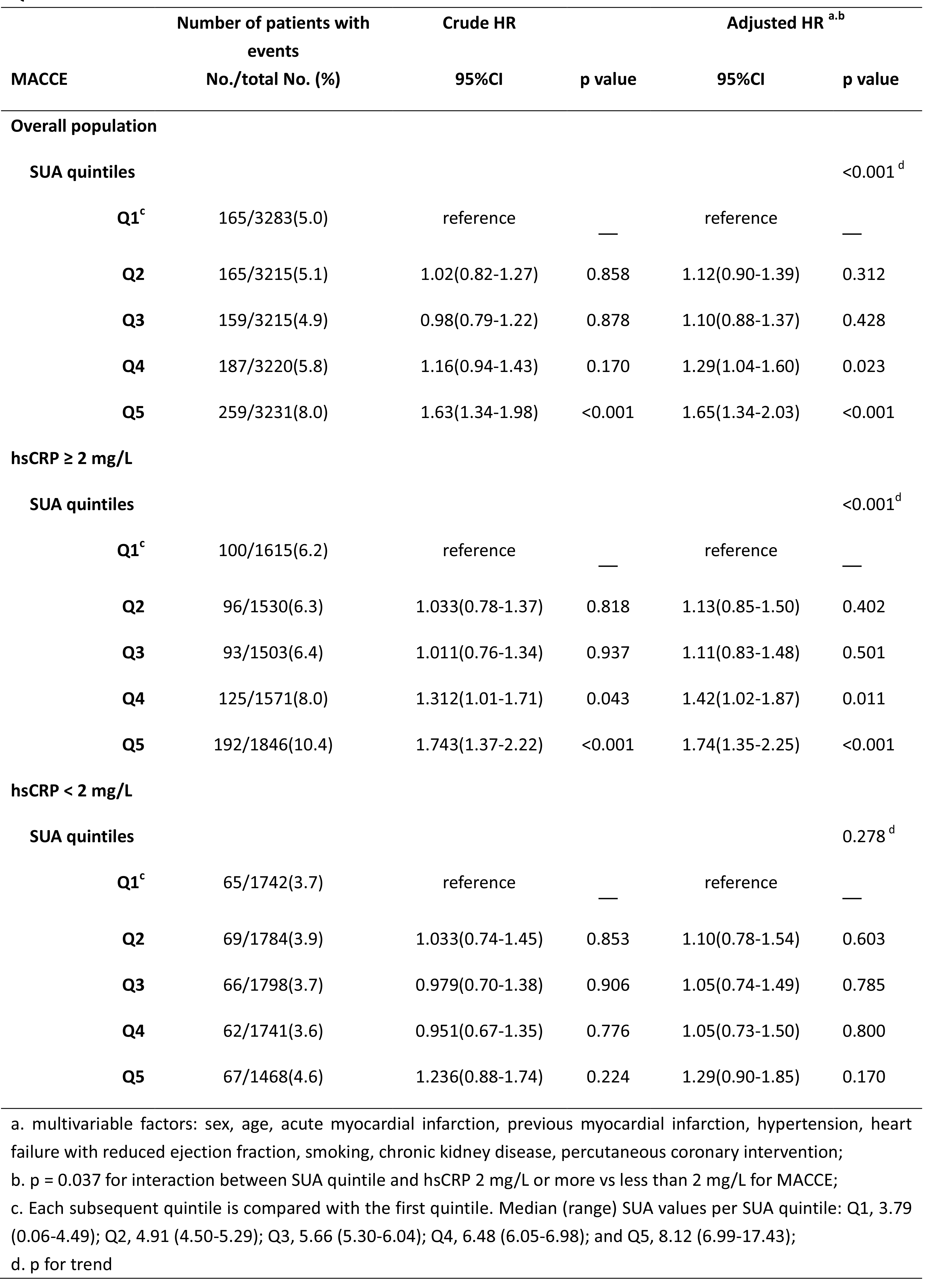
Multivariable analysis of association between MACCE according to achieved hsCRP Levels and SUA Quintile levels.

| Quintile levels |  |  |  |  |  |  |
| --- | --- | --- | --- | --- | --- | --- |
|  | Number of patients with events |  | Crude HR |  | Adjusted HR <sup>a,b</sup> |  |
| MACCE | No./total No. (%) |  | 95%CI | p value | 95%CI | p value |
| Overall population |  |  |  |  |  |  |
| SUA quintiles |  |  |  |  |  | <0.001 <sup>d</sup> |
| Q1<sup>c</sup> | 165/3283(5.0) |  | reference | — | reference | — |
| Q2 | 165/3215(5.1) |  | 1.02(0.82-1.27) | 0.858 | 1.12(0.90-1.39) | 0.312 |
| Q3 | 159/3215(4.9) |  | 0.98(0.79-1.22) | 0.878 | 1.10(0.88-1.37) | 0.428 |
| Q4 | 187/3220(5.8) |  | 1.16(0.94-1.43) | 0.170 | 1.29(1.04-1.60) | 0.023 |
| Q5 | 259/3231(8.0) |  | 1.63(1.34-1.98) | <0.001 | 1.65(1.34-2.03) | <0.001 |
| hsCRP ≥ 2 mg/L |  |  |  |  |  |  |
| SUA quintiles |  |  |  |  |  | <0.001 <sup>d</sup> |
| Q1<sup>c</sup> | 100/1615(6.2) |  | reference | — | reference | — |
| Q2 | 96/1530(6.3) |  | 1.033(0.78-1.37) | 0.818 | 1.13(0.85-1.50) | 0.402 |
| Q3 | 93/1503(6.4) |  | 1.011(0.76-1.34) | 0.937 | 1.11(0.83-1.48) | 0.501 |
| Q4 | 125/1571(8.0) |  | 1.312(1.01-1.71) | 0.043 | 1.42(1.02-1.87) | 0.011 |
| Q5 | 192/1846(10.4) |  | 1.743(1.37-2.22) | <0.001 | 1.74(1.35-2.25) | <0.001 |
| hsCRP < 2 mg/L |  |  |  |  |  |  |
| SUA quintiles |  |  |  |  |  | 0.278 <sup>d</sup> |
| Q1<sup>c</sup> | 65/1742(3.7) |  | reference | — | reference | — |
| Q2 | 69/1784(3.9) |  | 1.033(0.74-1.45) | 0.853 | 1.10(0.78-1.54) | 0.603 |
| Q3 | 66/1798(3.7) |  | 0.979(0.70-1.38) | 0.906 | 1.05(0.74-1.49) | 0.785 |
| Q4 | 62/1741(3.6) |  | 0.951(0.67-1.35) | 0.776 | 1.05(0.73-1.50) | 0.800 |
| Q5 | 67/1468(4.6) |  | 1.236(0.88-1.74) | 0.224 | 1.29(0.90-1.85) | 0.170 |

a. multivariable factors: sex, age, acute myocardial infarction, previous myocardial infarction, hypertension, heart failure with reduced ejection fraction, smoking, chronic kidney disease, percutaneous coronary intervention;
b. p = 0.037 for interaction between SUA quintile and hsCRP 2 mg/L or more vs less than 2 mg/L for MACCE;
c. Each subsequent quintile is compared with the first quintile. Median (range) SUA values per SUA quintile: Q1, 3.79 (0.06-4.49); Q2, 4.91 (4.50-5.29); Q3, 5.66 (5.30-6.04); Q4, 6.48 (6.05-6.98); and Q5, 8.12 (6.99-17.43);
d. p for trend

### Subgroup analysis

Further Subgroup analysis were conducted between groups of age, sex, whether PCI was performed, heart failure with reduced ejection fraction, acute myocardial infarction, diabetes mellitus, chronic kidney disease and baseline LDL-C levels. After fully adjusted multivariable analysis, except for HFrEF subgroup with hsCRP ≥ 2mg/L, the association of SUA quintiles and 2 years MACCE were consistent across subgroups (p for interaction > 0.05). In the setting of hsCRP ≥ 2mg/L, higher SUA levels were similarly associated with higher risk of MACCE in patients underwent PCI, with acute myocardial infarction, diabetes mellitus and baseline LDL-C less than 1.4mmol/L (Figure 4).

**Figure 4.**
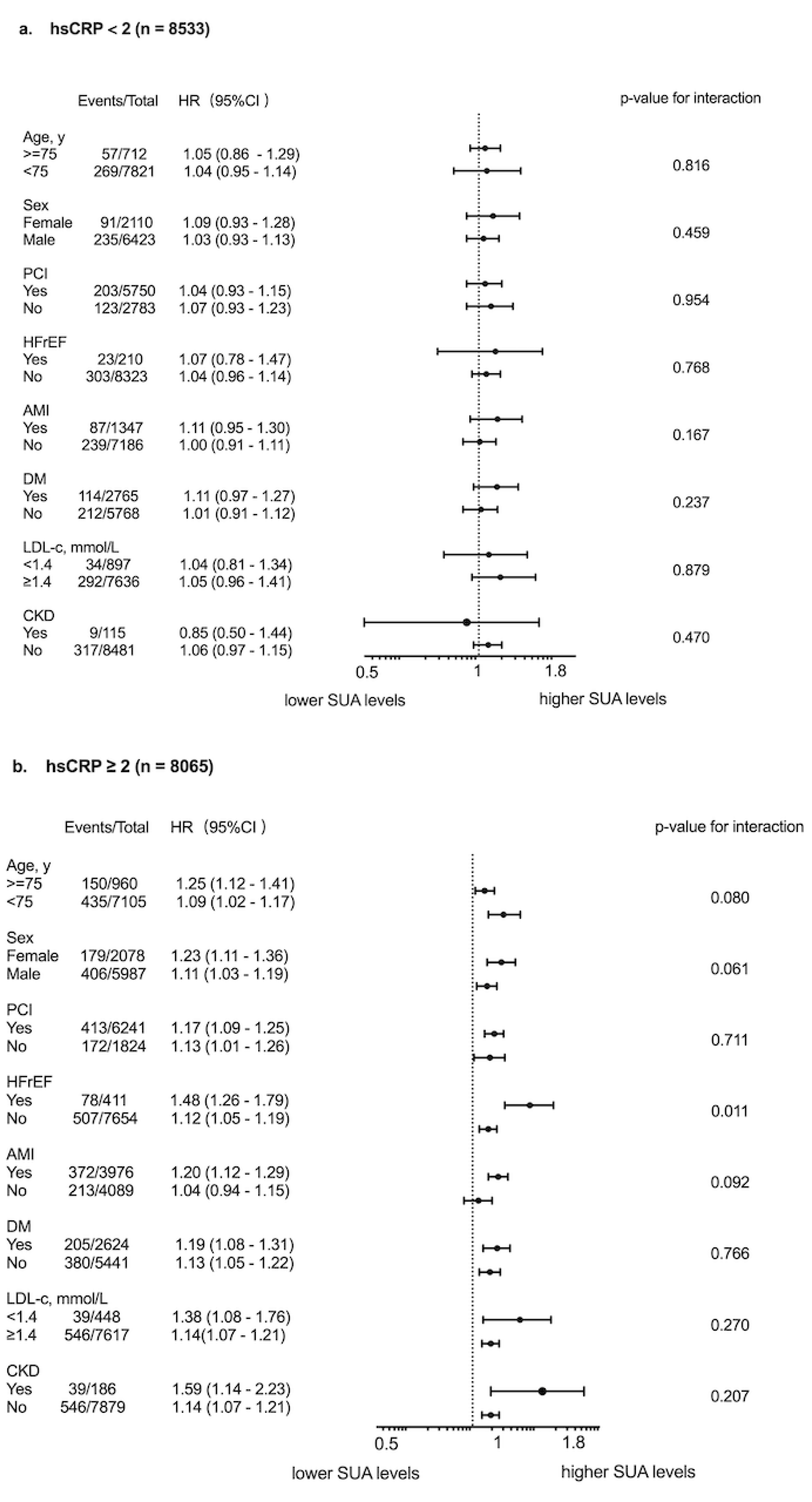
Subgroup analysis of major adverse cardiovascular and cerebrovascular events according to high-sensitivity C-reactive protein (hsCRP) and serum uric acid (SUA) levels in a Fully Adjusted Multivariable Model *multivariable model adjusted for sex, age, acute myocardial infarction, previous myocardial infarction, hypertension, heart failure with reduced ejection fraction, smoking, chronic kidney disease, percutaneous coronary intervention

## Discussion

To our knowledge, we demonstrated for the first time that SUA-associated cardiometabolic risk might be mediated by hsCRP levels, a marker for systemic inflammation, in a large multicenter cohort of CAD patients. The major findings of the current study are as follows: 1) After fully adjusted multivariable analysis, positive correlation of SUA levels and increased MACCE risk was only found in patients with greater degrees of systemic inflammation (hsCRP ≥ 2mg/L); 2) Each unit increase of SUA levels was associated with 11.3% increased risk of MACCE only when hsCRP level ≥ 2mg/L; 3) The association of SUA levels and 2 years MACCE were consistent across subgroups including age, sex, whether PCI was performed, acute myocardial infarction, diabetes mellitus, chronic kidney disease and baseline LDL-C levels less than 1.4mmol/L or not. These findings might elucidate the interdependence effect of the two known mediators on ASCVD risk: SUA and systemic inflammation.

Uric acid is the end-product of purine metabolism, which is mainly regulated by xanthine oxidoreductase (XOR), converting hypoxanthine to xanthine and finally to uric acid [17]. Hyperuricemia is closely associated with gout, chronic kidney disease and hypertension [2, 18], which are closely related to atherosclerosis. Recently, accumulating evidence demonstrated that uric acid may play significant roles in pathogenesis of cardiovascular disease and cardiovascular death [4, 5]. The underline pathogenesis of uric acid related cardiovascular risk is complicated, however oxidative stress and inflammation induced by uric acid are the main mechanism [9]. It is known that uric acid itself is chemically an important antioxidant in the extracelluar space. However, in the intracellular space, uric acid acts as a pro-oxidant agent, which induces endothelial dysfunction by intracellular oxidative stress and leads inflammation in vascular endothelial and smooth muscle cells [19, 20].

Inflammation has already been known to play an important role in the pathophysiology of atherogenesis and ischemic events [21]. Sufficient experimental and clinical evidence implicates that inflammation provide a series of pathways that link traditional risk factors, such as lipid and hypertension, to atherosclerosis [22, 23]. The biomarker hsCRP, which stands for systematic inflammation, is validated usefully for predicting both atherosclerotic and ischemic risk [24, 25]. Recent clinical trials have showed that therapy targeted on inflammation can reduce cardiovascular events in patients already received guideline recommended standard therapy. The “Canakinumab Anti-inflammatory Thrombosis Outcomes Study” (CANTOS) enrolled 10,061 patients with previous myocardial infarction and a hsCRP level of 2 mg/L or more despite of standard medical therapy. The participants were allocated randomly to receive three doses of canakinumab, an antibody that neutralizes the proinflammatory cytokine IL-1β. In patients who received 300-mg canakinumab every 3 months, the median level of hsCRP was reduced 41 percent greater than that with placebo, and the rate of recurrent cardiovascular events were significantly lowered beyond the effect of lipid lowering [10]. The results of the CANTOS trial affirmed that residual inflammation risk assessed by hsCRP levels was associated with future recurrent cardiovascular risk.

Previous study of in vitro human vascular cells had demonstrated that through stimulating expression of CRP, UA altered proliferation/migration of smooth muscle cell and nitric oxide release of endothelial cells [19] in the pathogenesis of UA-associated atherosclerosis. In this study, we reported in patients with hsCRP level ≥ 2mg/L, each unit increase of SUA levels was associated with 11.3% increased risk of MACCE. This finding provided clinical evidence that greater degree of systemic inflammation was associated with adverse SUA-induced cardiovascular risk. However, we also found a large portion of patients with high SUA levels but low systemic inflammation, in whom SUA levels were not associated with adverse prognosis. This finding implied that although SUA mediated CRP expression of vascular cells, more complex underlying mechanism of systemic inflammation in vivo might play important role in SUA-induced cardiovascular risk.

In general, SUA levels are higher in male than female for estrogen protection [26]. However, whether SUA-associated cardiovascular risk is different between female and male is still controversial. Sun et al [27] reported SUA levels were significantly associated with coronary atherosclerosis evaluated by 256-detector-row coronary computed tomographic angiography in female but not male. But Ando et al [28] found there was no difference in SUA-associated lipid-rich plaques by intravascular ultrasound between genders. In the current study, our subgroup analysis supported the consistency of SUA-associated cardiovascular risk in female and male. This finding suggested that when it comes to SUA related cardiovascular risk, systemic inflammation but not gender was the important factor that we should pay more attention to.

The improvement of cardiovascular outcomes of urate-lowering therapy (ULT) is still controversial. Allopurinol and febuxostat are both clinical commonly used ULT drugs reducing UA by inhibition of xanthine oxidase. Singh et al [29] reported that longer allopurinol use duration (at least 181days) was associated with reduced myocardial infarction in 29,298 elderly patients. Although with a relatively large sample size, the observational design was its major limitation. Subsequent small sample size randomized trials demonstrated benefit of ULT on surrogate endpoints such as blood pressure, endothelial function and carotid intima-media [30–32]. A recent systematic review including 6,458 participants from 28 trials with 506 major adverse cardiovascular events reported that ULT did not produce benefits on clinical outcomes including major adverse cardiovascular events [33]. Thus, lowering SUA alone to improve cardiovascular outcomes may not be effective. Our study reveals that SUA-associated cardiovascular risk appears to be significantly mediated by systemic inflammation, suggesting a potential benefit of combined ULT and anti-inflammation therapy in patients with greater systemic inflammation.

## Limitation

There are a number of limitations in the current study. First, the current analysis of the study was based on data from a large multicenter cohort. The observational design may preclude any definitive conclusion. Second, only baseline SUA and hsCRP levels were recorded in the current study and the conclusion focused on in-hospital SUA levels and inflammation status. With no follow-up results on SUA and hsCRP levels available, the effect of systemic inflammation and SUA trajectory related cardiovascular risk were still unclear. Third, two years follow-up duration was comparatively short for evaluating long-term outcomes. Finally, our study categorized participants into SUA quartiles and did not produce a certain cut-off value, which could hamper its clinical applicability.

## Conclusion

The present study demonstrates in a large multicenter prospective cohort that SUA-associated MACCE risk appears to be mediated by concomitant levels of systemic inflammation. This finding suggests a potential benefit of combined ULT and anti-inflammation therapy in patients with hyperuricemia and greater systemic inflammation.

## Author contributions

Jinqing Yuan, Yaling Han Xueyan Zhao, Jue Chen, Runlin Gao, Lei Song Participated in the study design. Ying Song, Weiting Cai, Lin Jiang, Jingjing Xu, Yi Yao, Na Xu, Xiaozeng Wang, Zhenyu Liu, Zheng Zhang, Yongzhen Zhang, Xiaogang Guo, Zhifang Wang, Yingqing Feng, Qingsheng Wang, Jianxin Li Participated in data analysis. Ying Song Wrote the manuscript. All authors participated in data interpretation and critical revision of the manuscript.

## Ethics approval and consent to participate

The Institutional Review Board approved the study protocol, which strictly adheres to the tenets of the Declaration of Helsinki.and the patients provided written informed consent before the intervention.

## Consent for publication

Written informed consent for publication was obtained from all participants.

## Data availability statement

Due to the nature of this research, participants of this study did not agree for their data to be shared publicly, so supporting data is not available.

## Acknowledgements

We thank all staff members for data collection, data entry, and monitoring as part of this study.

